# Therapeutic effects of nurse-led telephone follow-up on depression, anxiety, and stress in cardiovascular disease patients: a randomized clinical trial

**DOI:** 10.64898/2026.05.18.26353531

**Authors:** Hossein Mohsenipouya, Maryam Mahtabi, Fateme Yagoubi, Abolfazl Hosseinnataj, Raha Jafari Ghaleh, Taís Carpes Lanes

## Abstract

**Background:** Depression and anxiety are common among patients with cardiovascular disease (CVD) and are associated with poorer clinical outcomes, including increased complications and recurrent events. Nurse-led telephone follow-up after discharge has shown benefits in patient support and symptom management; however, its impact on psychological outcomes remains unclear. Therefore, this study aimed to evaluate the effects of a nurse-led telephone follow-up program on depression, anxiety, and stress levels in patients with cardiovascular disease.

**Methods:** An experimental study was conducted with 60 randomly selected patients from the Coronary Care Unit (CCU) department of a hospital in Iran, who were divided into two groups: an intervention group and a control group. The educational intervention was administered within two weeks after discharge. Data were collected via the Depression Anxiety Stress Scale (DASS-21). Descriptive analysis, Mann-Whitney and Wilcoxon tests, Generalized Estimating Equations (GEE) regression, and Spearman’s correlation were used for data analysis.

**Results:** The mean age of the patients was 57.43 ± 15.33 years. No significant differences were observed between the intervention and control groups regarding depression, anxiety, or stress scores (p>0.05). However, depression and anxiety scores decreased over time in both groups, with mean reductions of 1.53 and 1.18 points, respectively. Furthermore, an increase in patients’ ejection fraction (EF) was associated with a 0.1-point decrease in both depression and anxiety levels. No significant association was found between stress and the studied variables.

**Conclusions:** Depression and anxiety decreased over time in patients with cardiovascular disease. However, no significant differences were found between the intervention and control groups. Improved cardiac function (ejection fraction) was associated with lower depression and anxiety, highlighting the importance of integrating psychological and clinical care. Further studies with larger samples and longer follow-up are recommended.

## 1 Introduction

Depression and anxiety conditions affect a vast segment of the global population, with an estimated 300 million individuals suffering from depression and a similar number experiencing anxiety conditions, encompassing approximately 4–5% of the world’s population [1]. The coexistence of cardiovascular disease (CVD) and mental health disorders poses a significant and multidimensional challenge within the healthcare field. Understanding the intricate relationship between CVD and prevalent mental health conditions, such as depression and anxiety, is crucial for developing effective management strategies and optimizing patient outcomes. Extensive evidence suggests that exposure to daily stress or severe psychological trauma can significantly increase the risk of cardiovascular diseases[2, 3]. CVD accounts for approximately 46% of all deaths in Iran, making it the foremost health challenge in the nation[4]. In 2015, CVD was responsible for approximately 1 million DALYs, and projections suggest that this number could increase to nearly 1.73 million DALYs by 2025, primarily because of an aging population[5].

Depression commonly afflicts patients with CVD and is closely associated with CVD-related complications and increased healthcare costs. Notably, one out of every five patients with coronary artery disease or heart failure experiences depression, representing a prevalence three times higher than that of the general population [6]. CVD patients with depressive symptoms also face an increased risk of recurrent cardiovascular events and mortality [7]. Furthermore, prospective meta-analyses have revealed a 50% increased risk of developing and succumbing to CVD in individuals experiencing social isolation and loneliness, whereas work-related stress has also been linked to a 40% increased risk [8]. Additionally, qualitative studies have underscored patients’ belief that daily stress constitutes a pivotal underlying cause of CVD and associated risk factors, such as poor diet and sedentary lifestyle [9].

Anxiety, characterized by temporary fear, uncertainty, and apprehension about the future, represents another psychological facet with varying frequencies and intensities across individuals. Despite comparatively limited research on the relationship between anxiety and cardiovascular disorders compared with that between mental stress and depression [10], a meta-analysis revealed a greater risk of cardiovascular disease in otherwise healthy individuals with high anxiety levels [11]. Numerous factors have been suggested to elucidate the influence of anxiety on the onset or progression of CVD. These include the associations between anxiety and unhealthy behaviors such as smoking, excessive alcohol consumption, insufficient physical activity, and poor dietary choices, all of which contribute to an increased risk of cardiovascular disease[12, 13].

Postdischarge follow-up care is crucial for preventing disease exacerbation and improving patient outcomes. Telephone follow-up (TFU), often conducted by nurses, has proven effective in supporting patients after discharge by providing education and symptom management and enhancing communication with healthcare providers[14, 15]. A systematic review and meta-analysis highlighted the positive impact of TFU on cancer patients undergoing therapy, including reduced readmission rates, better medication adherence, and improved patient satisfaction[16]. These findings support the integration of TFU into standard care practices, particularly for patients with cardiovascular disease (CVD)[15].

While various studies have examined the role of telephone follow-up in reducing rehospitalization and promoting medication adherence while mitigating disease complications [15, 17, 18], few studies have explored the impact of TFU on stress, anxiety, and depression among cardiovascular patients, particularly after discharge [19, 20]. Hence, the primary objective of this study was to evaluate the effects of implementing the “nurse telephone follow-up” project on depression, anxiety and stress levels among cardiovascular patients. By employing a randomized controlled trial (RCT) design, this study ensures rigorous methodology and enhances the significance of the findings. The outcomes of this study will offer valuable insights into the efficacy of nurse-led telephone follow-up interventions in addressing mental health concerns and preventing readmissions among cardiovascular patients.

### Specific Objectives

1. To evaluate the effect of a nurse-led telephone follow-up intervention on depression levels among patients with cardiovascular disease after hospital discharge.

2. To assess the impact of the intervention on anxiety levels in patients with cardiovascular disease.

3. To determine the effect of nurse-led telephone follow-up on stress levels among cardiovascular patients following discharge.

## 2 Methods

### 2.1 Trial Design

This study was designed as a parallel-group, randomized controlled trial with an allocation ratio of 1:1. The trial followed a superiority framework, aiming to determine whether a nurse-led telephone follow-up intervention is superior to usual care in reducing depression, anxiety, and stress levels among patients with cardiovascular disease after hospital discharge.

No modifications were made to the trial design, eligibility criteria, outcomes, or analysis plan after the commencement of the study. All procedures were carried out exactly as prespecified in the approved protocol.

### 2.3 Trial Setting

This randomized controlled trial was conducted in a specialized hospital located in the eastern region of Mazandaran Province, Iran. The study setting included the inpatient cardiology ward and the outpatient post-discharge follow-up clinic of the hospital. This center functions as a tertiary referral hospital, providing advanced cardiovascular care to patients from Mazandaran and neighbouring provinces.

### 2.3 Sampling and Participants

This randomized controlled trial included patients with cardiovascular disease who were consecutively recruited from the hospital described above between November 23 and December 21, 2023. After providing informed consent and confirming eligibility, participants were randomly allocated to either the intervention group or the control group. All enrolled participants were followed for two weeks after hospital discharge, and the final follow-up assessments were completed by January 4, 2024. Baseline demographic characteristics, including age, gender, literacy level, and employment status, were collected prior to the intervention to assess the comparability of the study groups. The trial was completed as planned and was not stopped early.

### 2.4 Sample size calculation

The sample size was calculated based on the study by Shojai et al., in which the mean anxiety score was 11.05±7.68 in the intervention group and 18.76±11.79 in the control group [21]. Using a two-sided type I error rate of 5%, a statistical power of 80%, and the standard formula for comparing two independent means, the required sample size was estimated to be 27 participants per group. To account for an anticipated 10% dropout rate, the final sample size was increased to 30 participants in each group. The flow of participant allocation is presented in Figure 1. No interim analyses were planned or conducted, and no stopping guidelines were specified.

**Figure 1:**
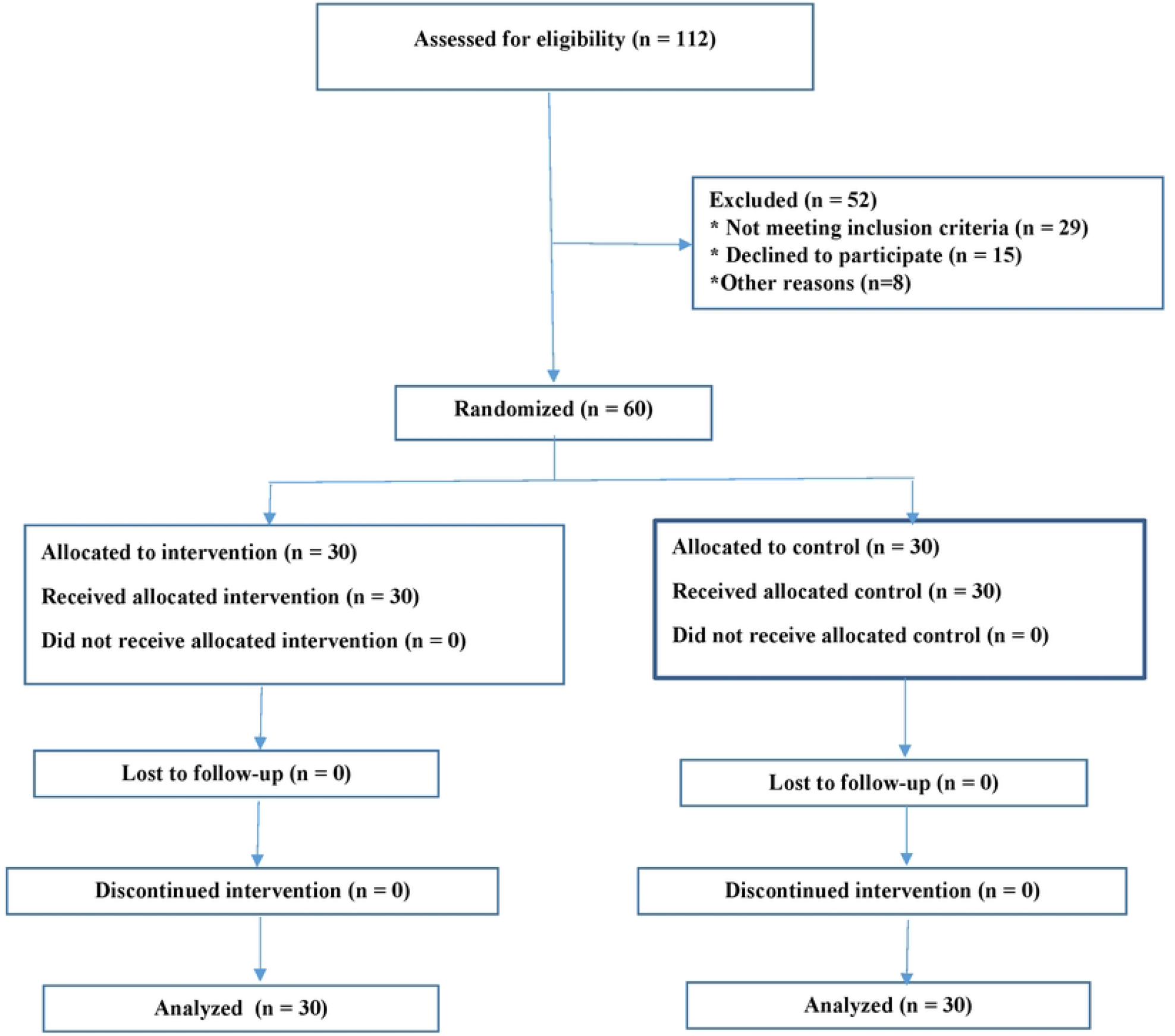
Flow of participants into the control and intervention groups

### 2.5 Randomisation

The random allocation sequence was generated by an independent statistician using a computer-generated list of random numbers. Simple randomization without restrictions was used to assign participants to either the intervention or control group.

### 2.6 Allocation Concealment

To prevent allocation bias, the random allocation sequence was kept by an independent researcher, and the individuals responsible for enrolling participants had no access to it. Group assignments were implemented using sequentially numbered, opaque, sealed, and light-proof envelopes. Each envelope was opened only after the participant had been enrolled and baseline data had been recorded, ensuring that the assigned group could not be predicted or altered beforehand.

### 2.7 Blinding

Blinding was not feasible due to the nature of the educational intervention.

### 2.8 Patient and Public Involvement

Patients or members of the public were not involved in the design, conduct, reporting, or dissemination plans of this trial. The research questions, outcome measures, and intervention procedures were developed entirely by the research team based on the existing literature and clinical guidelines.

### 2.4 Eligibility criteria for participants

The inclusion criteria for this study were: (a) willingness of both the patient and a family member to provide informed consent, (b) a confirmed diagnosis of cardiovascular disease by a specialist physician, © absence of any documented history of psychological disorders based on the patient’s medical records, and (d) no prior participation in similar educational or tele-follow-up programs.

Exclusion criteria were nonresponse to the nurse’s follow-up calls and failure to complete the study questionnaires. These criteria were established to ensure the recruitment of a homogeneous and appropriate sample capable of generating valid and reliable findings.

### 2.5 Eligibility criteria for sites and intervention providers

The study was conducted in a specialized hospital located in the eastern region of Mazandaran Province, Iran. The intervention was delivered by a trained nurse working in the cardiology department with clinical experience in cardiovascular patient care. The nurse received prior instruction regarding the telephone follow-up protocol and the procedures for providing patient education and support during the study period.

### 2.6 Intervention and Control Group

#### 2.6.1. Intervention

Participants in the intervention group received a structured telephone follow-up delivered by a trained cardiac nurse. At the time of discharge, each participant was provided with a written call schedule. A single follow-up call was conducted exactly two weeks after discharge and lasted approximately 10– 15 minutes. The call followed a standardized protocol and included assessment of the patient’s current clinical condition and symptoms, review of medication adherence and potential adverse reactions, education on fluid management, daily weight monitoring, and adherence to a heart-healthy diet, recommendations for safe physical activity and monitoring of vital signs, psychological support related to stress, anxiety, and emotional adjustment after cardiac hospitalization, smoking cessation counseling for eligible patients, and responding to any questions raised by patients or their caregivers. Educational content was based on national cardiovascular rehabilitation recommendations and was tailored to each patient’s level of awareness and psychosocial context. All calls were conducted by the same nurse to ensure consistency. The full intervention script and call checklist are available from the corresponding author upon reasonable request.

#### 2.6.2. Control Group

Participants in the control group received usual care, which included routine discharge education provided by the hospital nursing staff. No additional telephone follow-up, counseling, or structured educational support was delivered during the two-week post-discharge period. To ensure ethical compliance, participants in the control group received the same educational content provided to the intervention group only after the completion of the follow-up DASS-21 assessment.

During the study period, participants in both groups continued to receive standard medical care and pharmacological treatment as prescribed by their cardiologists. No additional structured educational or behavioral interventions were provided apart from the study intervention.

### 2.7 Outcomes

The pre-specified primary outcomes of this trial were depression, anxiety, and stress, assessed using the Depression Anxiety Stress Scale-21 (DASS-21). Each outcome was measured as a continuous variable using the relevant DASS-21 subscale score, with possible scores ranging from 0 to 42; higher scores indicated greater psychological distress.

Outcome assessments were conducted at two time points: baseline, prior to hospital discharge, and two weeks after discharge. The primary outcome evaluation focused on changes in depression, anxiety, and stress over time and differences between the intervention and control groups following the intervention. To further examine the effect of time and adjust for potential confounding variables, generalized estimating equation (GEE) regression analysis was performed.

For descriptive reporting, continuous variables were summarized using mean, standard deviation, and median, whereas categorical variables were presented as frequency and percentage.

### 2.8 Harms

No adverse events were expected due to the low-risk nature of the nurse-led telephone follow-up intervention. Harms were monitored non-systematically; participants were encouraged to report any discomfort or concerns during or after the telephone contact. No harms or unintended effects were reported in either group throughout the study period.

### 2.9 Measurements

#### 2.9.1 Sociodemographic and health-related variables

This study collected basic sociodemographic characteristics, including age, sex, educational degree (bachelor’s degree, master’s degree, doctorate), marital status, employment status, type of disease, underlying disease, ejection fraction level, hospitalization duration (day), and readmission status.

These factors were considered to provide a comprehensive understanding of the participants’ backgrounds and ensure a well-rounded analysis of the data.

#### 2.9.2 Depression Anxiety Stress Scale-21 (DASS-21)

The Depression Anxiety Stress Scale-21 (DASS-21) is a validated screening instrument used to assess mental health status[22]. It employs a four-point Likert scale, where participants indicate the degree to which specific statements apply to them. The DASS-21 consists of three subscales: depression, anxiety, and stress. Each subscale comprises seven items designed to measure different aspects of mental well-being. The depression subscale evaluates feelings of hopelessness, diminished interest or pleasure, and a negative outlook on life. The anxiety subscale assesses fear-related emotions and measures situational anxiety and autonomic arousal. The stress subscale measures difficulty in relaxation, tension, irritability, impatience, and other stress-related symptoms.

Scores on the DASS-21 for each subscale range from 0--42, with higher scores indicating a higher level of emotional distress [23].

The DASS-21 has demonstrated high reliability and validity in both clinical and nonclinical settings. Cronbach’s alphas for the subscales range from 0.87--0.94, indicating strong internal consistency [23]. The Cronbach’s alpha coefficient was acceptable for anxiety (0.79), stress (0.91), and depression (0.93) in the Persian version of the DASS-21[1]. This instrument has been widely used to evaluate mental health status, including during the COVID-19 pandemic, and has proven to be a valuable tool for assessing psychological well-being [24, 25]. In this study, the Cronbach’s alpha coefficient for the depression domain was 0.9, whereas for the anxiety and stress dimensions, it was 0.87.

### 2.10 Ethical considerations

This study strictly adhered to ethical guidelines and followed the principles outlined in the Declaration of Helsinki. Ethical approval for the study was obtained from the Ethics Committee of Mazandaran

University of Medical Sciences (Reference: IR.MAZUMS.REC.1401.269). Before proceeding with data collection, all participants were fully informed about the study’s objectives, potential risks, benefits, and other relevant information. Informed consent was obtained from all the subjects and their legal guardian(s). They were provided a comprehensive explanation to ensure their understanding of the study’s purpose and procedures. Importantly, participants were assured of their voluntary participation and the right to withdraw from the study at any point without consequences.

### 2.11 Statistical methods

In this study, various statistical methods were employed to analyse and interpret the data. Descriptive statistics, including the mean, standard deviation, median, frequency, and percentage, were used to summarize the variables under investigation. The Kolmogorov–Smirnov test was used to assess the normality of the data distribution, and the results indicated that the assumption of normality was not met. Therefore, non-parametric tests were applied.

The Mann–Whitney U test and chi-square test were used to compare demographic and clinical characteristics between the intervention and control groups. To evaluate changes in depression, anxiety, and stress scores, the Mann–Whitney U test was used to compare differences between the two groups, and the Wilcoxon signed-rank test was used to assess within-group changes before and after the intervention.

Generalized estimating equation (GEE) regression analysis was performed to examine the effect of time and to control for potential confounding variables. In addition, Spearman’s correlation coefficient was calculated to evaluate the relationships between depressions, anxiety, and stress scores.

All randomized participants were included in the analysis. Missing data were handled using available case analysis. All statistical analyses were performed using SPSS version 22. A p-value of less than 0.05 was considered statistically significant.

## 3 RESULTS

### 3.1 Participant Flow

A total of 112 participants were assessed for eligibility, of whom 60 were randomized into the control and intervention groups. The flow of participants throughout the trial is presented in Figure 1.

### 3.2 Baseline Characteristics

Among the participants, 55% were female and 86.7% were married. The mean age of the patients was 57.43 ± 15.33 years. The baseline demographic and clinical characteristics of participants in the intervention and control groups are presented in Table 1. The Mann–Whitney and chi-square tests were used to compare the demographic and clinical variables between the two groups. A significant difference was observed between the groups in terms of ejection fraction (EF) level (p = 0.036).

**Table 1.** Baseline demographic and clinical characteristics of participants in the intervention and control groups.

| Variables | Total Sample | Type of the participants |  |  |
| --- | --- | --- | --- | --- |
|  | N; (%) | Intervention; N (%) | Control; N (%) | p value |
| Gender |  |  |  |  |
| Male | 27 (45.0) | 14 (46.7) | 13 (43.3) | 0.795 |
| Female | 33 (55.0) | 16 (53.3) | 17 (56.7) |  |
| Marital status |  |  |  |  |
| Single | 8 (13.3) | 5 (16.7) | 3 (10) | 0.706 |
| Married | 52 (86.7) | 25 (83.3) | 27 (90) |  |
| Education level |  |  |  |  |
| Illiterate | 17 (28.4) | 8 (26.7) | 9 (30.0) | 0.208 |
| Elementary | 9 (15.0) | 7 (23.3) | 2 (6.7) |  |
| Guidance | 6 (10.0) | 4 (13.3) | 2 (6.7) |  |
| Diploma | 26 (43.3) | 11 (36.7) | 15 (50.0) |  |
| Masters | 2 (3.3) | 0 | 2 (6.7) |  |
| Employment status |  |  |  |  |
| Free | 14 (23.3) | 9 (30.0) | 5 (16.6) | 0.450 |
| Employee | 3 (5.0) | 1 (3.3) | 2 (6.7) |  |
| Farmer | 2 (3.3) | 2 (6.7) | 0 |  |
| Military | 8 (13.3) | 3 (10) | 5 (16.7) |  |
| Other | 33 (55.0) | 15 (50) | 18 (60.0) |  |
| Types of diseases |  |  |  |  |
| Diabetes | 3 (5.0) | 2 (6.7) | 1 (3.3) | 0.513 |
| Heart disease | 47 (78.3) | 24 (80.0) | 23 (76.7) |  |
| HTN | 4 (6.7) | 3 (10.0) | 1 (3.3) |  |
| Pulmonary edema | 1 (1.7) | 1 (3.3) | 0 |  |
| LOC | 1 (1.7) | 0 | 1 (3.3) |  |
| Skin disease | 2 (3.3) | 0 | 2 (6.7) |  |
| Renal disease | 1 (1.7) | 0 | 1 (3.3) |  |
| Aneurysm | 1 (1.7) | 0 | 1 (3.3) |  |
| Hospitalization frequency |  |  |  |  |
| 0 | 42 (70.0) | 18 (60.0) | 24 (80.0) | 0.052 |
| 1 | 12 (20.0) | 9 (30.0) | 3 (10.0) |  |
| 2 | 2 (3.3) | 0 | 2 (6.7) |  |
| 3 | 2 (3.3) | 1 (3.3) | 1 (3.3) |  |
| 5 | 2 (3.3) | 2 (6.7) | 0 |  |
| Underlying disease |  |  |  |  |
| Yes | 51 (85.0) | 24 (80.0) | 27 (90.0) | 0.472 |
| No | 9 (15.0) | 6 (20.0) | 3 (10.0) |  |
| Variables | Mean ± SD | Mean ± SD | Mean ± SD | p value<sup>+</sup> |
| Age | 57.43±15.33 | 55.97±14.64 | 58.90±16.10 | 0.455 |
| Hospitalization duration (day) | 4.33±3.68 | 3.80±2.04 | 4.87±4.77 | 0.416 |
| Ejection Fraction level | 40.42±14.35 | 36.17±15.68 | 44.67±11.67 | 0.036 |
| *: Chi-square test +: Mann–Whitney test |  |  |  |  |

### 3.3 Changes in Depression, Anxiety, and Stress Scores

Table 2 presents the mean scores of depression, anxiety, and stress in the intervention and control groups before and after the intervention. Within-group analysis showed significant reductions in depression and anxiety scores in the intervention group (p = 0.007 and p = 0.011, respectively) and in the control group (p < 0.001 and p = 0.001, respectively). However, no significant differences were observed between the intervention and control groups at either measurement time for depression, anxiety, or stress scores (all p > 0.05).

**Table 2.** Comparison of DASS-21 depression, anxiety, and stress scores between the intervention and control groups before and after the intervention.

| Outcome | Time | Intervention (n = 30) |  | Control(n = 30) |  | p value* |
| --- | --- | --- | --- | --- | --- | --- |
|  |  | Mean (SD) | Mean Rank | Mean (SD) | Mean Rank |  |
| Depression | Before | 7.63 (5.42) | 32.42 | 4.67 (5.15) | 28.58 | 0.394 |
|  | After | 6.40 (5.20) | 33.18 | 4.63 (3.51) | 27.82 | 0.232 |
|  | p value<sup>+</sup> | 0.007 |  | <0.001 |  |  |
| Anxiety | Before | 8.57 (5.14) | 31.80 | 7.50 (4.49) | 29.20 | 0.563 |
|  | After | 7.57 (4.17) | 33.35 | 6.13 (3.98) | 27.65 | 0.204 |
|  | *p* value+ | 0.011 |  | 0.001 |  |  |
| Stress | Before | 9.30 (5.57) | 31.08 | 8.63 (3.92) | 29.92 | 0.795 |
|  | After | 8.97 (5.15) | 30.40 | 9.73 (7.47) | 30.60 | 0.964 |
|  | *p* value+ | 0.361 |  | 0.423 |  |  |

\* *Mann–Whitney U test (between-group comparison)*
+ *Wilcoxon signed-rank test (within-group comparison)*

**Table 3.** Factors affecting depression, anxiety and stress scores via GEE regression.

| Variables | Depression |  | Anxiety |  | Stress |  |
| --- | --- | --- | --- | --- | --- | --- |
|  | B (95% CI) | *p* value | B (95% CI) | *p* value | B (95% CI) | *p* value |
| **Next time** | -1.53 (-2.1, -0.9) | **<0.001** | -1.18 (-1.7, -0.6) | **<0.001** | 0.38 (-0.7, 1.4) | 0.478 |
| **Group 1** | 0.56 (-1.7, 2.9) | 0.635 | 0.38 (-1.6, 2.3) | 0.700 | -0.94 (-3.9, 2.0) | 0.532 |
| **EF level** | -0.10 (-0.2, 0.02) | **0.021** | -0.10 (-0.2, -0.03) | **0.007** | -0.11 (-0.2, 0.1) | 0.066 |

*B = regression coefficient; CI = confidence interval; EF = ejection fraction.*
*Group 1, intervention group;*
*Reference group: control group.*
*Time indicates assessment two weeks after the intervention.*
*GEE model with Gaussian distribution, identity link, and exchangeable correlation structure.*

### 3.4 Factors Associated with Depression, Anxiety, and Stress

The relationships between time, EF level, and psychological outcomes were examined using the GEE regression model. The mean depression score was 1.53 points lower after the intervention compared with before the intervention (p < 0.001). Additionally, each unit increase in EF was associated with a 0.1 decrease in the depression score (p = 0.021). Similarly, the mean anxiety score decreased by 1.18 points after the intervention (p < 0.001), and higher EF levels were associated with lower anxiety scores (p = 0.007). No significant associations were found between stress scores and the studied variables.

### 3.5 Correlation between Depression, Anxiety, and Stress

Table 4 presents the correlations between depression, anxiety, and stress scores before the intervention. Significant positive correlations were observed among all three variables, indicating that higher scores in one psychological domain were associated with higher scores in the others. These findings suggest a strong interrelationship among depression, anxiety, and stress prior to the intervention.

**Table 4.** Correlations between depression, anxiety, and stress scores before the intervention.

| Variables | Depression | Anxiety | Stress |
| --- | --- | --- | --- |
| Depression | 1 |  |  |
| Anxiety | 0.87 | 1 |  |
| Stress | 0.77 | 0.85 | 1 |

Table 5 presents the correlations between depression, anxiety, and stress scores after the intervention. Similar to the pre-intervention findings, significant positive correlations were found among all three variables, demonstrating that increases in one score were associated with increases in the other scores. These results indicate that the interrelationship among depression, anxiety, and stress remained consistent after the intervention.

**Table 5.** Correlations between depression, anxiety, and stress scores after the intervention.

| Variables | Depression | Anxiety | Stress |
| --- | --- | --- | --- |
| Depression | 1 |  |  |
| Anxiety | 0.88 | 1 |  |
| Stress | 0.78 | 0.82 | 1 |

### 3.6 Harms

No harms, adverse events, or unintended effects were reported in either the intervention group or the control group during the study period.

## 4 Discussion

### 4.1 Interpretation of findings

This study aimed to assess the impact of an educational intervention, specifically telephone follow-up (TFU), on depression, anxiety, and stress levels among cardiovascular disease (CVD) patients before and after discharge. Previous studies have supported the effectiveness of intervention programs in reducing depression, anxiety, and stress. For example, Shoushi et al. reported significant improvements in depression, stress, and anxiety scores in a family support program for patients undergoing open-heart surgery [26]. Aktas et al. reported lower depression scores in the training group than in the standard care group at discharge [27]. Another study reported significant differences in depression, anxiety, and pain among intervention groups [28].

Before the intervention was implemented, this study revealed no significant difference between the groups in terms of suffering from mental health problems, indicating appropriate randomization of both groups. After the nurses conducted the TFU, it was expected that the intervention group would report less severe psychiatric problems than would the control group. Although the mean scores of mental health problems decreased after the TFU intervention in the treatment group, the difference compared with those in the control group was not significant. Similarly, Missler et al. conducted a study to evaluate the efficacy of a brief educational, psychological intervention in preventing postpartum parental stress and reducing symptoms of depression and anxiety [29]. Surprisingly, both the intervention and control groups demonstrated increased postpartum stress, and no significant group differences were found in parenting stress or mental health outcomes, suggesting that the universal prevention program did not effectively alleviate parental anxiety or enhance the quality of care [29]. Several factors may have contributed to the nonsignificant outcome of the TFU intervention in our study. The relatively short duration between the pre- and postintervention assessments may have limited the intervention’s ability to induce substantial changes in depression, anxiety, and stress levels.

Additionally, the modest sample size might have influenced the study’s statistical power, potentially affecting the ability to detect significant differences between groups.

However, in this study, the mean scores of depression and anxiety appeared to decrease after TFU in both the intervention and control groups. These findings suggest that implementing a two-week educational program holds promise for reducing anxiety and depression among patients with cardiovascular diseases. However, a decrease in anxiety and depression was also observed in the control group, indicating that the presence of other factors contributed to the reduction. While the educational program had some positive effects, it is crucial to consider the broader context of patient care in relation to anxiety and depression. Mental health outcomes are influenced by various individual and contextual factors, including personal coping strategies, social support networks, and individual resilience [30-34]. Future research should explore the multifaceted nature of patient experiences and identify additional interventions or strategies to alleviate mental health outcomes in a home-based setting. This may involve incorporating other psychological interventions, enhancing social support systems, or utilizing alternative treatment approaches to optimize patient outcomes[34-36]. It is also important to consider the individual variability in patient responses to the educational program. Some patients may benefit more from the intervention than others do on the basis of their unique circumstances and needs. Understanding these individual differences can help tailor interventions to maximize their effectiveness.

In this study, an intriguing observation was made regarding the relationship between ejection fraction (EF) levels and the severity of psychiatric problems. For each one-point increase in the EF level, there was a corresponding decrease of 0.1 in depression and anxiety levels. These findings are consistent with previous studies, which reported significantly lower EF percentages in patients with depression than in those without the disease [37]. For example, one study reported that patients with reduced ejection fraction (HFrEF) presented higher anxiety and depression scores than did those with preserved ejection fraction (HFpEF) and non-HF controls[38]. These findings suggest that psychological distress is prevalent in individuals with HF, with depression affecting up to 60% of patients[39]. Furthermore, patients with coronary artery disease, particularly those classified as NYHA Class III/IV and experiencing unstable angina, presented higher rates of anxiety and depressive symptoms during unstable disease periods than did those with stable angina [40]. These findings suggest that anxiety and depression scores decreased over time in both groups. However, because no significant differences were observed between the groups, the specific effect of the educational program should be interpreted with caution.

### 4.2 Implications

- **Integration of telephone follow-up (TFU):** Although nurse-led TFU tended to reduce depression and anxiety among cardiovascular patients, the lack of significant differences from the control group suggests that TFU should be integrated as part of a broader, multidimensional support system for patients postdischarge.
- **Continuous Mental Health Monitoring:** The observed reductions in anxiety and depression scores—even in the control group—underscore the need for ongoing mental health screening and support for all cardiac patients, regardless of the specific intervention received.
- **Personalized patient support:** The relationship observed between ejection fraction (EF) and psychological outcomes suggests that mental health interventions may need to be tailored on the basis of individual cardiac function and risk factors, making personalized care plans especially important.
- **Interdisciplinary and Family Involvement:** Leveraging family support and interdisciplinary collaboration among healthcare providers may increase the efficacy of psychological interventions and provide a more holistic approach to patient management.
- **Research and evaluation:** Short intervention periods and modest sample sizes may limit observable changes; therefore, further large-scale, longitudinal studies are necessary to establish best practices for psychological support in cardiovascular care.
- **Policy development:** Policymakers should consider supporting the implementation of accessible mental health interventions—such as TFU—as part of standard postdischarge care, accompanied by education for both patients and their caregivers.

### 4.3 Limitations

This study has several limitations. First, the relatively small sample size may have reduced the statistical power to detect significant differences between the intervention and control groups. Second, participants were recruited from a single hospital, which may limit the generalizability of the findings to other populations and healthcare settings. Third, the follow-up period was relatively short and may not have been sufficient to evaluate the long-term effects of the intervention on depression, anxiety, and stress. In addition, psychological outcomes were measured using self-reported questionnaires, which may be subject to reporting bias. Finally, multiple statistical analyses were performed, which may increase the possibility of type I error. Future multicenter studies with larger sample sizes and longer follow-up periods are recommended to confirm these findings and further evaluate the effectiveness of telephone follow-up interventions in patients with cardiovascular disease.

### 4.4 Conclusion

This study suggests that a two-week educational program implemented for patients with CVD holds promise in reducing anxiety and depression. Nonetheless, these findings underscore the importance of considering factors beyond educational interventions in impacting patients’ mental health outcomes, particularly within the realm of remote follow-up care. Subsequent research endeavours should delve deeper into investigating the efficacy of home-based interventions and the influence of the hospital environment on shaping mental health outcomes.

## Data Availability

The minimal dataset underlying the findings of this study, including anonymized demographic information and questionnaire scores (DASS?21), is available in a public data repository. All relevant data have been uploaded to the Mendeley Data repository and can be accessed via the following DOI: https://data.mendeley.com/datasets/x45xrhwmzh/1. No identifying personal information is included in the dataset.

## Author Declarations

### Data Availability

The datasets generated and analyzed during the current study are available from the corresponding author upon reasonable request. The trial protocol and statistical analysis plan are provided as Supplementary Information files accompanying this article.

## Acknowledgments

The authors would like to extend their heartfelt gratitude and appreciation to the participants of this study. Their participation is invaluable.

